# *Clostridioides difficile* PCR Positivity on Successive Days after Treatment Initiation

**DOI:** 10.1101/2022.08.30.22279403

**Authors:** Nabin K. Shrestha, Jeffrey Shu, Nishan K. Shrestha, Steven M. Gordon

## Abstract

Among 1282 patients with a positive *Clostridioides difficile* PCR test who were retested within 14 days, the proportion of positive tests rapidly decreased on successive days within the first week since initiation of treatment. Between days 7 and 14 after treatment initiation, approximately 25% of tests remained positive.

## INTRODUCTION

There is a general recommendation against testing for cure of *C. difficile* infection by PCR, based on an assertion that tests remain positive in 60% of patients after successful treatment, thereby implying that repeat testing provides no value [1,2]. However, data supporting this assertion (and the assumption that repeated testing offers limited clinical value) are scarce. One commonly cited study with 76 patients with *C. difficile* infection, found that 6 (55%) of 11 patients treated with vancomycin had detectable *C. difficile* in their stools (by culture) 10-15 days after treatment [3]. This has been cited to support the recommendation against repeat testing, and the recommendation repeated so many times over many years that most people believe it to be based on sound science. Misrepresentation of study findings has also contributed to this belief. One widely cited study, which found that patients with toxin-positive *C. difficile* infection had a milder course of illness, and better outcomes, than toxin-negative *C. difficile* infection, was misleadingly titled, “Overdiagnosis of *Clostridium difficile* infection in the molecular test era” [4].

*Clostridioides difficile* PCR is one of the most commonly conducted PCR tests in our hospital. A positive test usually, correctly or incorrectly, leads to initiation of antimicrobial therapy. Although repeat *C. difficile* PCR testing within 7 days is actively discouraged, particularly following a positive test, some patients nevertheless have additional tests done within a few days of a prior test. This circumstance provides an opportunity to explore whether PCR tests actually remain positive in the majority of patients after initiation of treatment.

## METHODS

All patients who underwent *C. difficile* PCR testing at Cleveland Clinic between January 1, 2011 and January 1, 2022, were identified from the enterprise data vault. Those who had a positive test, and were tested at least once more within the subsequent 14 days, were included. For these patients, initial positive tests followed by tests done within 14 days of the positive test, were selected for analysis. The study was approved by the Cleveland Clinic Institutional Review Board as exempt human subject research, with a waiver of informed consent and HIPPAA authorization (IRB no. 22-563).

For each included patient, the date of the first positive test (index test) was considered day zero. Subsequent tests for different patients occurred on various days following day zero, until day 14. On each of the days from days 1 through 14, the proportions of all tests done that day that were positive, were calculated. Ninety-five percent confidence intervals were determined using 10,000 bootstrapped samples. A polynomial regression line was fitted to percentage positive against day. The analysis was done using R version 4.1.2 [5].

## RESULTS

Of 123, 596 patients who were tested at least once by PCR for *C. difficile* infection, 1282 met our inclusion criteria. Their ages were distributed with a mean of 62 and standard deviation of 20 years, 60% were female, and the index positive tests were distributed across all years of the study, with each individual year contributing between 5 and 13% of all index positive tests.

The figure shows the proportion of tests positive on successive days following the initial positive test and a fitted line from a polynomial regression model. The regression model was: lm(formula = pos_proportion ∼ poly(relative_test_day, 3), data = data). The overall regression was statistically significant (adjusted R^2^ 0.92, F(3, 11) = 52.9, p-value < 0.001). Over 14 days since presumed initiation of treatment, the proportion positive decreased within a day of starting treatment, and dropped with each successive day over the first week of treatment to a plateau around 25%.

## DISCUSSION

This study calls into question the assumption that *C. difficile* PCR tests remain positive in the majority of patients after initiation of treatment. After starting effective treatment for any infection, the burden of living microorganisms is expected to decrease. Additionally, one would expect a living human body to recycle free amino acids and nucleotides with great efficiency. There should be no reason to expect dead microorganisms to be capable of protecting their proteins and nucleic acids from tissue proteases and nucleases. So, although PCR is capable of detecting dead as well as living microorganisms, it is unlikely that nucleic acid from dead microorganisms persists for long durations within a living human body. It is therefore likely that positive PCR test results in about 25% of patients more than a week after starting treatment are from the presence of living microorganisms. It is interesting that in clinical treatment trials, relapse within 28 days of completion of treatment ranged from 13 – 30% [6,7], which is remarkably similar to the proportion of tests that remain positive after one week of treatment.

One prior study from Mayo Clinic found that the median time to negative PCR in *C. difficile* infection was 9 days from treatment initiation [8]. This is not consistent with our study’s findings, but caution is warranted in generalization of the findings of that study to the general population, as only 50 patients were included in the study despite being an 8-year-long prospective study at a large tertiary referral care center. Moreover, the patients included in the study were sicker at baseline compared to what would be expected in the general population: 24% of patients had prior *C. difficile* infection and 24% had underlying inflammatory bowel disease.

Although our study is limited in that it represents only a very small fraction of all patients with *C. difficile* infection, it is unlikely that a higher proportion of patients who underwent repeat testing would have been expected to test negative than those who were not retested. Therefore, this study may have overestimated, but certainly did not underestimate, the proportion of *C. difficile* PCR tests that remain positive on any specific day after initiation of treatment. Although we did not collect treatment data for this study, practicing infectious disease physicians know all too well that a positive *C. difficile* PCR test generally prompts treatment for *C. difficile* infection, and our hospitals are no different in that regard. It can be safely presumed that almost all patients with a positive *C. difficile* PCR test in this study, were treated for *C. difficile* infection.

In light of our study’s findings, entirely dismissing repeat PCR testing in *C. difficile* infection appears inopportune, and it would be prudent to re-examine the clinical utility of testing for cure in *C. difficile* infection. The study on *C. difficile* PCR positivity on follow-up testing from Mayo Clinic did find that a positive PCR test during or after treatment completion was associated with a higher risk of recurrence [8]. We hypothesize that the correct interpretation of a positive PCR test for a microorganism in a living human is that it represents living microorganisms or microorganisms that were alive until very recently. If this is true, a positive *C. difficile* PCR test done a week or two after initiation of treatment should be a useful predictor of relapse of infection, a hypothesis that can be readily tested.

## Data Availability

All data produced in the present study are available upon reasonable request to the authors

## Notes

## Author contributions

N. K. S.: Conceptualization, methodology, validation, investigation, data curation, software, formal analysis, visualization, writing-original draft preparation, writing-reviewing and editing, supervision, project administration. J. S.: Data curation, software, validation, formal analysis, visualization, writing-reviewing and editing. N. K. S.: Data curation, validation, visualization, writing-reviewing and editing. S. M. G.: Project administration, resources, writing-reviewing and editing.

### Funding

None.

## FIGURES

**Figure 1.**
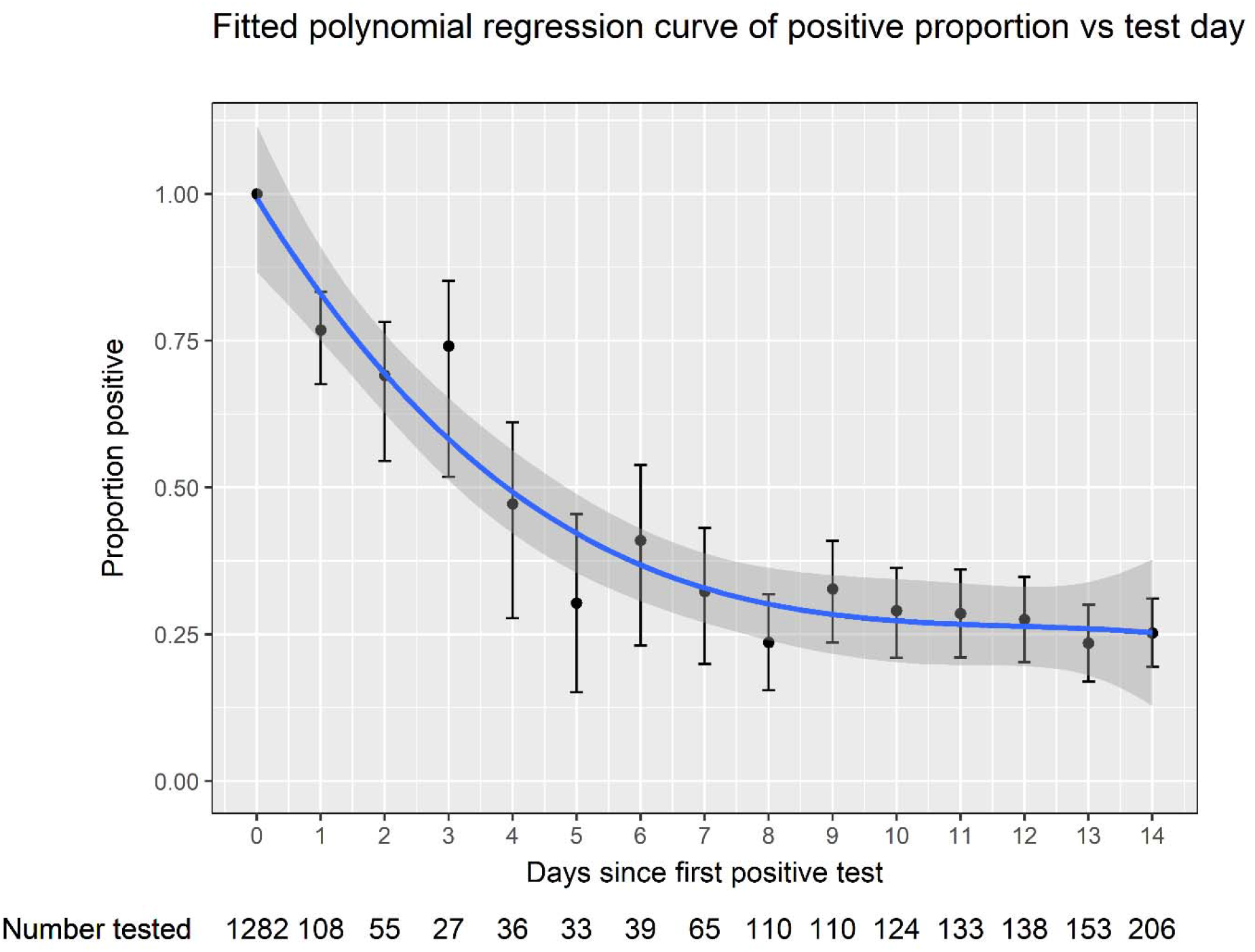
Proportions of *C. difficile* PCR tests that remained positive on successive days after the first positive test, with error bars representing 95% confidence intervals, along with a fitted polynomial curve to the proportions positive by day, with the shaded areas along the fitted line representing 95% confidence bands. The numbers below the x-axis represent the number of patients who were tested on that particular day relative to the index positive test.

## REFERENCES

1. Gupta A, Khanna S. Repeat Clostridium difficile Testing. JAMA 2016; 316:2422–2423.

2. McDonald LC, Gerding DN, Johnson S, et al. Clinical Practice Guidelines for Clostridium difficile Infection in Adults and Children: 2017 Update by the Infectious Diseases Society of America (IDSA) and Society for Healthcare Epidemiology of America (SHEA). Clin Infect Dis 2018; 66:e1– e48.

3. Abujamel T, Cadnum JL, Jury LA, et al. Defining the Vulnerable Period for Re-Establishment of Clostridium difficile Colonization after Treatment of C. difficile Infection with Oral Vancomycin or Metronidazole. PLOS ONE 2013; 8:e76269.

4. Polage CR, Gyorke CE, Kennedy MA, et al. Overdiagnosis of Clostridium difficile Infection in the Molecular Test Era. JAMA Intern Med 2015; 175:1792–1801.

5. R Core Team. R: A language and environment for statistical computing. 2021;

6. Louie TJ, Miller MA, Mullane KM, et al. Fidaxomicin versus Vancomycin for Clostridium difficile Infection. N Engl J Med 2011; 364:422–431.

7. Cornely OA, Crook DW, Esposito R, et al. Fidaxomicin versus vancomycin for infection with Clostridium difficile in Europe, Canada, and the USA: a double-blind, non-inferiority, randomised controlled trial. Lancet Infect Dis 2012; 12:281–289.

8. Saha S, Yadav D, Pardi R, Patel R, Khanna S, Pardi D. Kinetics of polymerase chain reaction positivity in patients with Clostridioides difficile infection. Ther Adv Gastroenterol 2021; 14:17562848211050444.

